# A comprehensive characterization of blood group antigen variants in the Middle Eastern population genomes - Insights into genetic epidemiology

**DOI:** 10.1101/2023.11.20.23298777

**Authors:** Mercy Rophina, Kavita Pandhare, Vinod Scaria

## Abstract

**Background:** The Middle Eastern population is characterized by increased prevalence of various Mendelian disorders owing to increased rates of consanguinity. Especially in disease conditions which require chronic transfusion support, it becomes important to know the blood group characteristics of potential donors to increase the likelihood of transfusion success. As there prevails a gap in knowledge about the population specific overall blood antigen profiles, this study seeks to utilize next generation sequencing datasets to unravel the comprehensive landscape of clinically significant minor blood group alleles in the middle eastern population.

**Methods:** This study utilizes the genetic variation data from a range of public datasets including the Greater Middle East Variome, the Qatar genome and exomes and the Iranome datasets to estimate the genotypic and phenotypic frequencies of blood group alleles in the Middle Easterners. The estimated frequencies were duly compared with major global populations to identify significant similarities or differences if any.

**Results:** A total of 77 unique ISBT approved blood group alleles were found commonly in all datasets. 8 variants (rs8176058, rs1058396, rs565898944, rs28362692, rs2071699, rs34783571, rs60322991 and rs57467915) belonging to KELL, KIDD, COLTON, H, JUNIOR and LANGEREIS blood groups were found clinically significant with previously reported evidence on transfusion complications. 730 variants were found to span exonic or splicing regions out of which 70 were predicted to be potentially deleterious by at least four computational tools.

**Conclusions:** This study serves first of its kind to extensively characterize the known and novel blood alleles in the Middle Easterners. A comprehensive user-friendly online resource named **alnasab - *Alleles and antigens in Arab and Persian populations associated with blood groups*** was also developed as a dependable reference for future transfusion research. The resource is accessible at https://clingen.igib.res.in/alnasab/

**Key points:**

- Large scale Middle Eastern population sequencing datasets including *The Greater Middle Eastern Variome, Genomes and exomes from Qatar and the Iranome* datasets were used in the study.
- A total of **2828** exomes and **88** genomes were analyzed accounting for a total of
- **18717** unique human blood group related variants.
- **2443** exonic variants were extracted which systematically included **1505** non-synonymous variants, **766** synonymous variants, **50** stopgain variants and **3** stop loss variants.
- Blood group associated variants identified in the study are provided as a comprehensive online repository - **alnasab,** *Alleles and antigens in Arab and Persian populations associated with blood groups.* The resource is accessible at https://clingen.igib.res.in/alnasab/

**Visual abstract:** 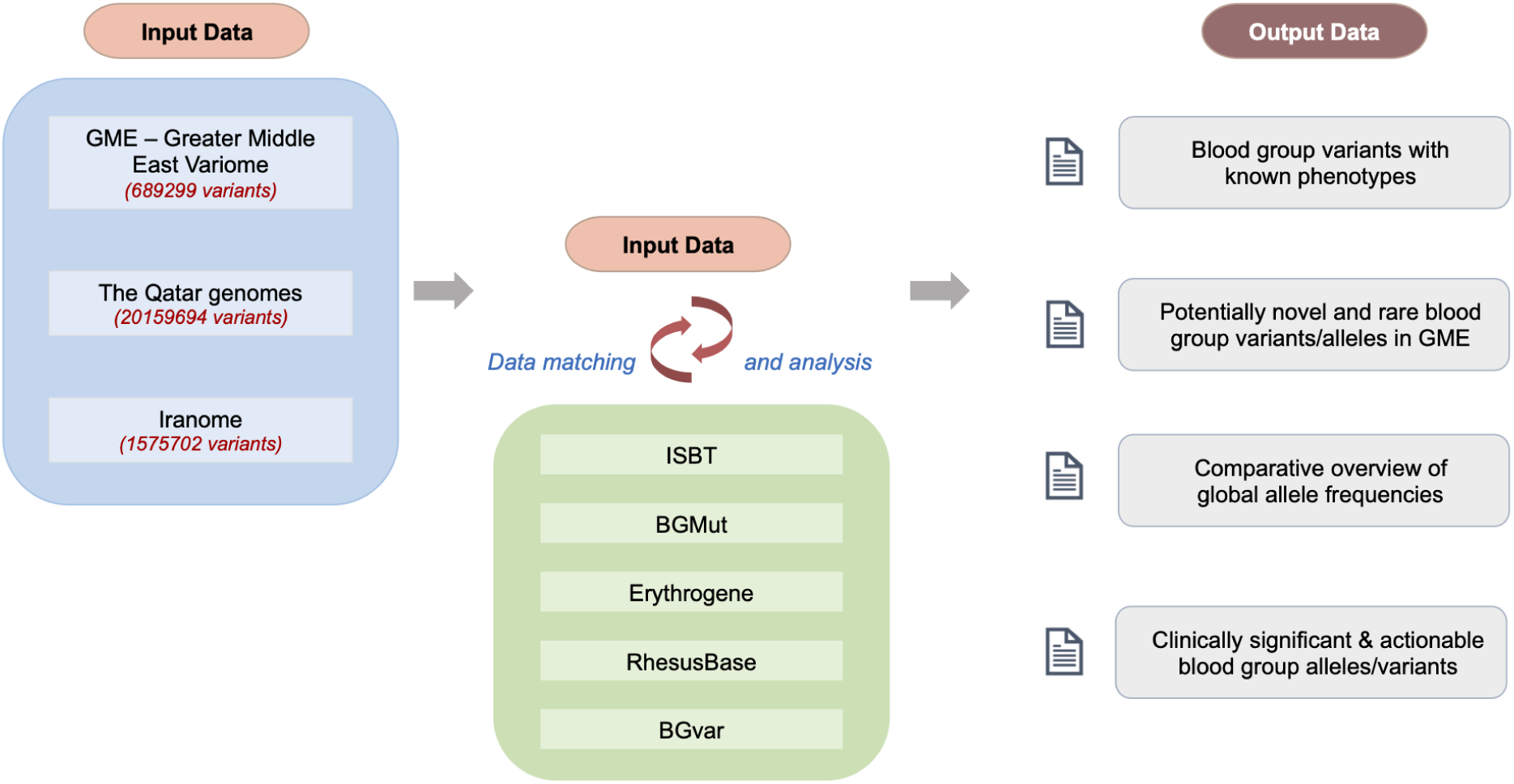

## Introduction

Blood group antigens are genetically encoded and currently encompass 44 blood group systems, encoding a total of 349 antigens mapping to 50 genes as recognized and approved by the International Society for Blood Transfusion (ISBT). Complications accompanying blood transfusion events are often induced by blood products which escape serological vigilance. Research over the past decade has widened the scope of genomics in immunohematology by highlighting the existence of geographical and ethnicity based differences in blood genotype and phenotype frequencies. (1), (2), (3).

The Middle Eastern region situated between Africa, Europe and South Asia is the abode of a population which has been central to our understanding of human evolution, history and migration (4), (5), (6), (7). Encompassing about 17 countries with an estimated population of around 411 million, this region has often remained underrepresented in large global population scale genomic studies. However, with a recent spurt in interest, various population scale sequencing projects have been undertaken aimed at understanding the genomic diversity and architecture of the Arab populations.(8), (9), (10), (11), (12).

Only a limited number of studies have investigated the prevalence of blood group antigens in Middle Eastern populations, primarily focusing on the ABO and RH systems along with few other clinically significant minor blood groups (13), (14), (15), (16), (17). However, these studies have been limited by their use of standard serological testing or medium throughput molecular typing techniques, and therefore there is a paucity of a comprehensive population-specific molecular blood group profile.

It is well established that genetic disorders are a significant burden in the Middle East owing to their increased rates of consanguinity. This has resulted in high prevalence of various autosomal recessive disorders. In particular, genetic blood disorders including Sickle cell disease (SCD) and 𝛽-thalassemia, the most common hemoglobinopathies have been reported to substantially contribute to the disease burden of this population (18), (19). A review estimated that the overall incidence and prevalence of SCD in Middle Eastern countries ranged from approximately 0.004% - 2.1% respectively (20). Such inherited blood disorders owing to their chronic nature tend to impose heavy medical, emotional and financial burden. Although the overall burden of such illnesses encompassing various medical aspects of the disease largely remain uncharacterized in Middle Eastern countries, conditions like thalassemia major (TM) and SCD demand long term transfusion therapy.

In recent years, there have been systematic efforts to determine the blood group phenotypes of regular blood donors visiting donation centers in the United Arab Emirates. Advancements in next generation sequencing techniques would undoubtedly enable the large population scale deployment of such efforts in the region. The availability of numerous public Middle Eastern genomic datasets encouraged us to computationally decipher the blood antigen profiles of Middle Easterners as previously described by Schoeman et al in 2019.

In this study we aim to compile a comprehensive collation of blood alleles prevailing in the middle eastern population along with a systematic comparison with other major population datasets including the 1000 Genomes Project and gnomAD. In addition, we developed a user-friendly online resource *alnasab* - *Alleles and antigens in Arab and Persian populations associated with blood groups* which is a comprehensive catalog of genetic variants associated with human blood group systems identified in Middle Easterners. The resource is accessible online at https://clingen.igib.res.in/alnasab/

## Materials and methods

### Population scale genomic datasets

Whole genome and whole exome sequencing datasets generated by population specific studies in the Greater Middle Eastern region were compiled and used for the analysis. This includes data from the Greater Middle East Variome (21), The Qatar genome and exomes (10) and the Iranome (9) genome projects. These accounted for a total of 2828 high quality exomes and 88 genomes (GME : 1111 exomes; Qatar : 88 genomes and 917 exomes ; Iranome : 800 exomes), encompassing 689299, 20159694 and 1575702 total genetic variants respectively. The population scale data is a collation of major ethnic groups of the middle eastern region including North Western Africa (NWA), North East Africa (NEA), Arabian Peninsula (AP), Turkish Peninsula (TP), Persia and Pakistan (PP), Syrian Desert (SD), Qatari subgroups and Iranian Arabs, Azeris, Baluchs, Kurds, Lurs, Turkmen and Persians. All the datasets correspond to human genome 19 (GRCh37/hg19) assembly. Genetic variations were retrieved from the variant call format (VCF) files and were used for further analyses.

### Human blood group related alleles and variations

Clinically approved genomic coordinates of 50 genes associated with 43 human blood group systems and 2 erythroid specific transcription factors were fetched from Locus Genomic Reference (LRG) (22) and were used for analysis as described previously (23). There exists a handful of public resources that provide comprehensive collection of human blood alleles and antigens including the International Society of Blood Transfusion (ISBT) (24), The Blood Group Antigen Gene Mutation Database (dbRBC) (25), Erythrogene (26) RhesusBase (27), Blood Antigens (28,29) and BGvar (30). A comprehensive list of approved and predicted human blood group alleles were retrieved in a pre-formatted template (30) and was used as the reference data for further allele matching procedures.

### Data processing and variant annotations

Primary analysis involved the filtering of all genetic variations spanning the LRG approved genomic coordinates of blood group genes and erythroid specific transcription factors. Subsequently, the filtered variants were systematically annotated for their functional consequences from a range of computational tools including SIFT (31), Polyphen (32), LRT, MutationTaster, Mutation Assessor (33), FATHMM (34), PROVEAN (35), CADD (36), GERP (37), PhyloP (38) and PhastCons using ANNOVAR (v. 2018-04-06) (39).

### Known and novel blood group alleles/variants identification

Secondary stages of variant analyses involved the filtering and identification of well known and novel blood group alleles. Reference dataset was used to match all the variants which already had a known associated blood phenotype and reference genome nomenclature was conferred to those blood group systems with no blood group associated variants. In addition, potentially novel and rare blood group variants were also filtered and analyzed. A variant was deemed potentially novel if it lacked previous literature reports and SNP identification number (dbSNP ID). Variants with minor allele frequencies (MAF) < 5% were also filtered and checked for their association with blood group related phenotypes. A complete schematic representation of the workflow is shown in **Figure 1**.

**Figure 1.**
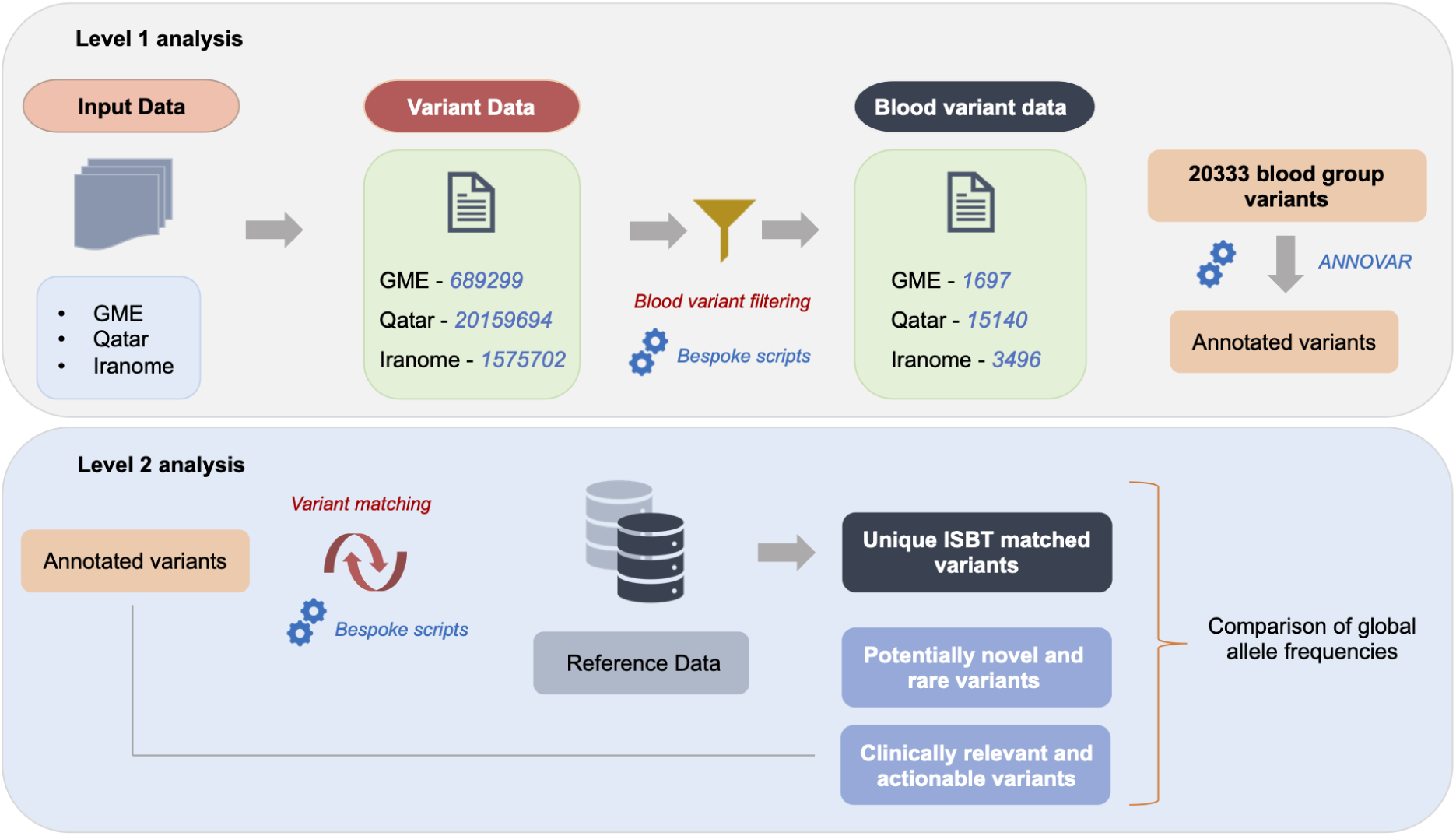
Schematic representation of the methodology followed for data analysis

### Prediction of blood phenotypes in Qatari subpopulations

The publicly available genomic data of 1005 Qatari individuals comprising 5 major subpopulations were analyzed in the study to decipher the pattern of distribution of blood phenotypes among the Qatari subpopulations. The variant call file was systematically subsetted into each subpopulation using VCFTOOLS (40). Bespoke scripts were used to generate the number of samples with homozygous and heterozygous genotypes. Similarities and distinct differences in blood alleles and phenotype frequencies were studied. Minor allele frequencies were compared among the subpopulations and statistical significance was observed using Fisher’s exact test with a p-value < 0.05.

### Estimation and systematic comparison of blood allele frequencies

Blood group associated variants filtered from all the datasets were systematically compiled in variant call or annotated formats along with their genotype information. Allele frequencies were estimated using PLINK {ref}. In addition, allele frequencies of the variants were fetched from major global population datasets including 1000 Genomes project {ref}, Exome Aggregation Consortium (ExAC v.0.3) {ref} and Genome Aggregation Database (gnomAD) {ref} and were used for comparison. Frequency of the filtered alleles among the subpopulations were also calculated.

### Identification of clinically significant and actionable blood group alleles

Variants with known blood group associated phenotypes filtered from the datasets were further checked for their clinical relevance in transfusion procedures and in pregnancy settings. Literature databases and public resources like Pubmed and Google scholar were systematically queried to retrieve relevant clinically significant evidence of alleles.

### Compendium of Greater Middle East blood group variants - Database architecture

The filtered variants were compiled into a preformatted mastersheet and annotations were duly mapped. The variant data and corresponding annotations were transformed to JavaScript Object Notation format and ported onto MongoDB 3.4.1. The user-friendly web interface for querying the database was coded in PHP 7.0, AngularJS, HTML, Bootstrap 4 and CSS. The web server was configured in Apache HTTP server.

## Results

### Overview of blood group alleles in Greater Middle East

We retrieved a total 18717 unique variants mapping to 50 human blood group genes, out of which 1697, 15140 and 3496 variants were from the Greater Middle East Variome, The Qatar genome and the Iranome datasets respectively. **Supplementary Table 1** provides a comprehensive summary of all the variant counts in blood group genes fetched from the above mentioned datasets. Of the variants, 2428 and 15 variants were found across exonic and splicing regions respectively. Of these, there were 1505 non synonymous SNVs, 766 synonymous SNVs, 50 stop gain and 3 stop loss variants. Variant summary and functional classifications across the datasets are schematically represented in **Figure 2**.

**Figure 2.**
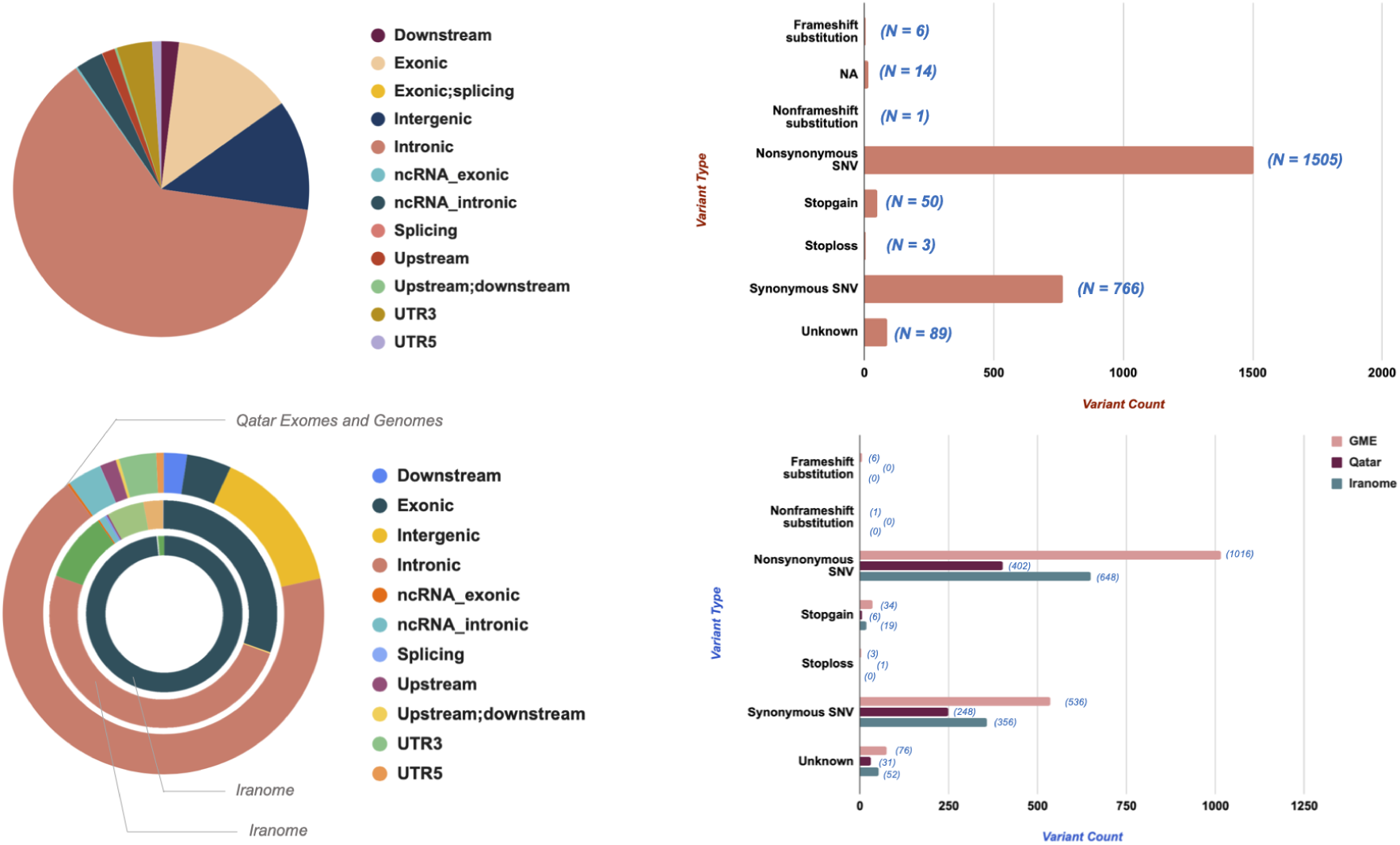
Overview of the functional classification of blood group related variants in various datasets used in the study

### Characterization of genetic variations in human blood group genes

Of the variants which mapped to the 50 blood group genes, a total of 147, 110 and 112 variants were previously shown to be associated with blood group related phenotypes in GME, Qatar and Iranome datasets respectively. Of these variants, a total of 77 unique variants belonging to 20 blood group systems were overlapping in all three datasets used in this study. The details of the variants along with their frequencies are tabulated in **Table 1**. **Figure 3** illustrates the distribution of these variants and their corresponding allele frequencies in various datasets. Forty-six (46) of the total 77 variants were SNVs mapping back to 16 blood groups and the rest were combination mutations for which blood phenotypes could not be predicted. Phenotypes were reported the same as that of the reference genome (hg19) nomenclature for the rest blood group systems as previously described (41), (23). **Table 2** summarizes the phenotypes of the variants and their corresponding zygosity information across the datasets. Phenotype frequencies of blood group systems with known blood group associated variants and those with reference genome nomenclature are detailed in **Supplementary Table 2.**

**Figure 3.**
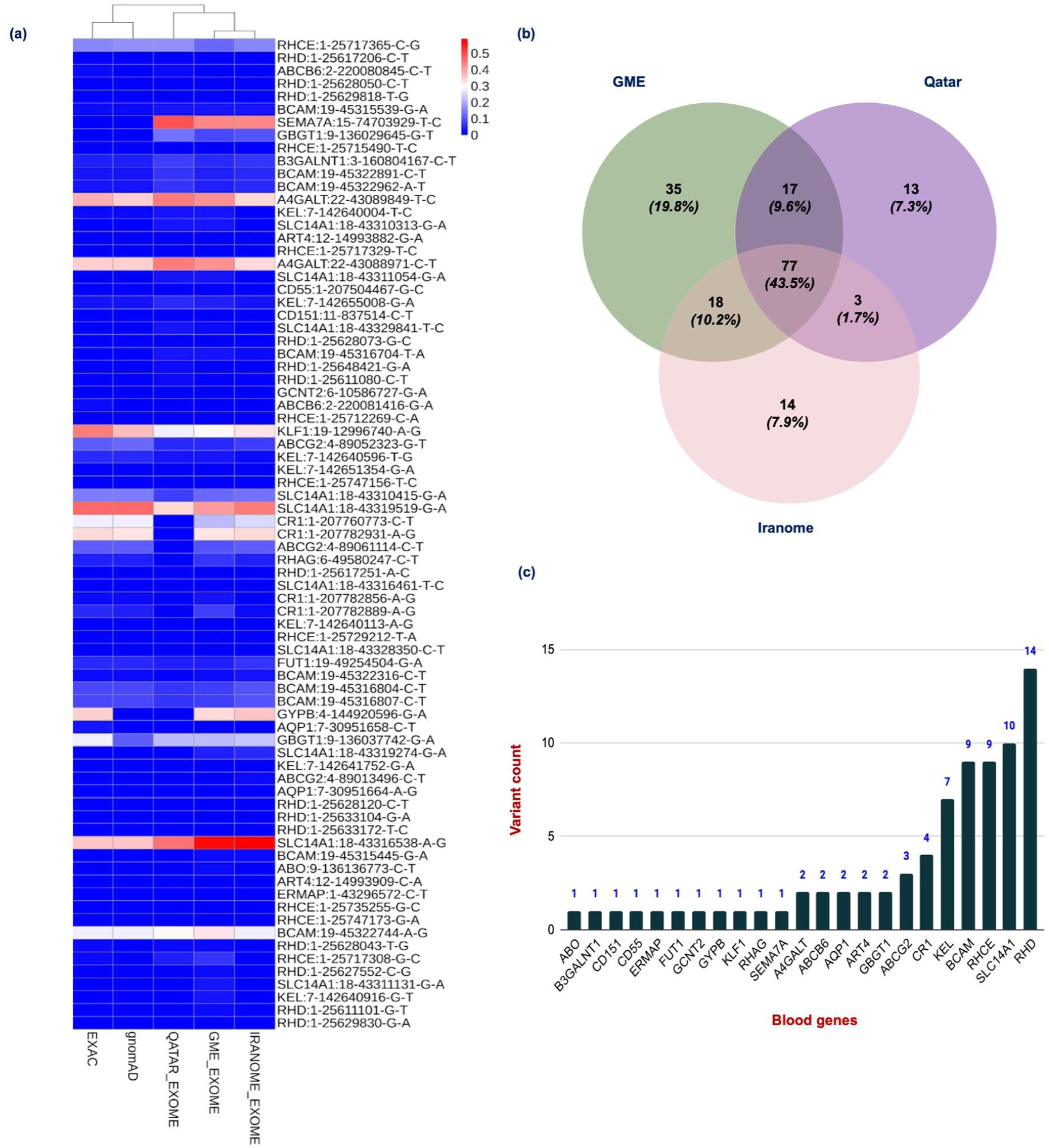
Schematic illustration of the distribution of common variants with blood group associated phenotypes along with their corresponding allele frequencies. (A), Heatmap illustrating the similarities and differences in the allele frequencies among various datasets (B), Distribution of blood group related variants in various datasets used in the study (C) Distribution of common alleles among human blood group systems

**Table 1.** Summary of variants with blood group associated phenotypes found commonly in all the Middle Eastern datasets used in the study.

| CHR | POS | RSID | REF | ALT | BLOOD<br>GROUP<br>SYSTEM | GENE<br>NAME | CATEGO<br>RY | GME_EX<br>OME_AF | QATAR_E<br>XOME_AF | IRANOME_<br>EXOME_AF | EXAC_<br>AF | gnomA<br>D_AF |
| --- | --- | --- | --- | --- | --- | --- | --- | --- | --- | --- | --- | --- |
| 11 | 837514 | rs139042921 | C | T | **RAPH** | **CD151** | Combinat<br>ion<br>Mutation<br>s | 0.00252<br>016129 | 0.0064 | 0.000625 | 0.0001<br>09 | 0.0000<br>88 |
| 1 | 207504467 | rs60822373 | G | C | **CROMER** | **CD55** | SNVs | 0.00453<br>1722054 | 0.006 | 0.00125 | 0 | 0 |
| 1 | 207760773 | rs3737002 | C | T | **KNOPS** | **CR1** | SNVs | 0.22205<br>43807 | 0 | 0.24875 | 0.2750<br>08 | 0.2824<br>56 |
| 1 | 207782856 | rs17047660 | A | G | **KNOPS** | **CR1** | SNVs | 0.02970<br>795569 | 0 | 0.0025 | 0.02141<br>1 | 0.01815<br>6 |
| 1 | 207782889 | rs17047661 | A | G | **KNOPS** | **CR1** | SNVs | 0.07552<br>870091 | 0 | 0.011875 | 0.0548<br>3 | 0.0460<br>45 |
| 1 | 207782931 | rs6691117 | A | G | **KNOPS** | **CR1** | SNVs | 0.32426<br>98892 | 0 | 0.33875 | 0.3341<br>07 | 0.3213<br>6 |
| 12 | 14993882 | rs28362798 | G | A | **DOMBROCK** | **ART4** | SNVs | 0.00805<br>6394763 | 0.007 | 0.001875 | 0 | 0 |
| 12 | 14993909 | rs28362797 | C | A | **DOMBROCK** | **ART4** | Combinat<br>ion<br>Mutation<br>s | 0.00604<br>2296073 | 0.0032 | 0.000625 | 0.0050<br>74 | 0.0039<br>38 |
| 1 | 25611080 | rs2301155 | C | T | **RH** | **RHD** | SNVs | 0.00753<br>4983854 | 0.0133 | 0.00721785 | 0.0007<br>44 | 0.0008<br>14 |
| 1 | 25611101 | rs199509194 | G | T | **RH** | **RHD** | Combinat<br>ion<br>Mutation<br>s | 0.01885<br>775862 | 0.0048 | 0.00197109 | 0.0066<br>53 | 0.0055<br>86 |
| 1 | 25617206 | rs113982491 | C | T | **RH** | **RHD** | SNVs | 0.00371<br>5498938 | 0.0059 | 0.0026560<br>4 | 0.0066<br>54 | 0.0057<br>81 |
| 1 | 25617251 | rs17418085 | A | C | **RH** | **RHD** | Combinat<br>ion<br>Mutation<br>s | 0.00641<br>7112299 | 0 | 0.0026809<br>7 | 0.0066<br>27 | 0.0054<br>44 |
| 1 | 25627552 | rs1053355 | C | G | **RH** | **RHD** | SNVs | 0.01397<br>849462 | 0 | 0.0053404<br>5 | 0.0050<br>9 | 0.0046<br>95 |
| 1 | 25628043 | rs1053356 | T | G | **RH** | **RHD** | SNVs | 0.02355<br>460385 | 0.0171 | 0.0060402<br>7 | 0.01211<br>4 | 0.0103<br>46 |
| 1 | 25628050 | rs148014996 | C | T | **RH** | **RHD** | Combinat<br>ion<br>Mutation<br>s | 0.00319<br>8294243 | 0.0064 | 0.00134048 | 0.0040<br>99 | 0.0034<br>47 |
| 1 | 25628073 | rs1053359 | G | C | **RH** | **RHD** | SNVs | 0.00373<br>5325507 | 0.0047 | 0.00201072 | 0 | 0 |
| 1 | 25628120 | rs1053362 | C | T | **RH** | **RHD** | SNVs | 0.00482<br>3151125 | 0.0021 | 0.0026738 | 0.0011<br>82 | 0.0010<br>57 |
| 1 | 25629818 | rs141833592 | T | G | **RH** | **RHD** | SNVs | 0.002212<br>389381 | 0.0065 | 0.00134953 | 0.0045<br>2 | 0.0033<br>3 |
| 1 | 25629830 | rs150606530 | G | A | **RH** | **RHD** | SNVs | 0.011615<br>04425 | 0.0041 | 0.0026845<br>6 | 0.0042<br>9 | 0.0035<br>32 |
| 1 | 25633104 | rs146292192 | G | A | **RH** | **RHD** | SNVs | 0.00423<br>7288136 | 0.0021 | 0.0026809<br>7 | 0.0005<br>72 | 0.0005<br>91 |
| 1 | 25633172 | rs138235491 | T | C | **RH** | **RHD** | SNVs | 0.00477<br>7070064 | 0.0021 | 0.00266312 | 0.0006<br>09 | 0.0006<br>39 |
| 1 | 25648421 | rs150059028 | G | A | **RH** | **RHD** | SNVs | 0.00826<br>9018743 | 0.0158 | 0.0087956<br>7 | 0.0041<br>84 | 0.0043<br>62 |
| 1 | 25712269 | rs116261244 | C | A | **RH** | **RHCE** | SNVs | 0.001007049345 | 0 | 0.00125 | 0.003571 | 0.002871 |
| 1 | 25715490 | rs1132765 | T | C | **RH** | **RHCE** | SNVs | 0 | 0.0016 | 0.001875 | 0.000512 | 0.000466 |
| 1 | 25717308 | rs1053361 | G | C | **RH** | **RHCE** | SNVs | 0.06344410876 | 0.0368 | 0.008125 | 0.022719 | 0 |
| 1 | 25717329 | rs144163296 | T | C | **RH** | **RHCE** | SNVs | 0.004028197382 | 0.0035 | 0.001875 | 0.001343 | 0.000951 |
| 1 | 25717365 | rs609320 | C | G | **RH** | **RHCE** | SNVs | 0.1299093656 | 0.1716 | 0.161875 | 0.162186 | 0.169006 |
| 1 | 25729212 | rs1053345 | T | A | **RH** | **RHCE** | SNVs | 0.0005035246727 | 0.0006 | 0.006875 | 0.001903 | 0.001786 |
| 1 | 25735255 | rs57992529 | G | C | **RH** | **RHCE** | SNVs | 0.005415162455 | 0.0022 | 0.000877193 | 0 | 0 |
| 1 | 25747156 | rs138268848 | T | C | **RH** | **RHCE** | SNVs | 0.008503401361 | 0.0041 | 0.00875 | 0.01081 | 0.010245 |
| 1 | 25747173 | rs142971926 | G | A | **RH** | **RHCE** | SNVs | 0.009029345372 | 0.0035 | 0.000625 | 0 | 0 |
| 15 | 74703929 | rs741761 | T | C | **JMH** | **SEMA7A** | Combination Mutations | 0.4310171198 | 0.4922 | 0.431875 | 0 | 0 |
| 18 | 43310313 | rs113578396 | G | A | **KIDD** | **SLC14A1** | Combination Mutations | 0.027693857 | 0.026 | 0.009375 | 0.006567 | 0 |
| 18 | 43310415 | rs2298720 | G | A | **KIDD** | **SLC14A1** | Combination Mutations | 0.1218529708 | 0.0796 | 0.1375 | 0.147459 | 0.151956 |
| 18 | 43311054 | rs113029149 | G | A | **KIDD** | **SLC14A1** | Combination Mutations | 0.02923387097 | 0.0299 | 0.009375 | 0.008122 | 0.00692 |
| 18 | 43311131 | rs28994287 | G | A | **KIDD** | **SLC14A1** | Combination Mutations | 0.02947154472 | 0 | 0.009375 | 0.007885 | 0.006644 |
| 18 | 43316461 | rs9948825 | T | C | **KIDD** | **SLC14A1** | SNVs | 0.001510574018 | 0 | 0.000625 | 0.001507 | 0.001153 |
| 18 | 43316538 | rs2298718 | A | G | **KIDD** | **SLC14A1** | Combination Mutations | 0.5730110775 | 0.4562 | 0.590625 | 0.36427 | 0.361846 |
| 18 | 43319274 | rs17675299 | G | A | **KIDD** | **SLC14A1** | SNVs | 0.03675730111 | 0 | 0.053125 | 0 | 0 |
| 18 | 43319519 | rs1058396 | G | A | **KIDD** | **SLC14A1** | SNVs | 0.4058408862 | 0.3323 | 0.439375 | 0.46954 | 0.471467 |
| 18 | 43328350 | rs565898944 | C | T | **KIDD** | **SLC14A1** | SNVs | 0 | 0 | 0.0025 | 0.000651 | 0.00066 |
| 18 | 43329841 | rs28898897 | T | C | **KIDD** | **SLC14A1** | Combination Mutations | 0.01762336354 | 0.0264 | 0.010625 | 0.004547 | 0.004667 |
| 19 | 12996740 | rs2072597 | A | G | **KLF1** | **KLF1** | Combination Mutations | 0.2962771458 | 0.2835 | 0.325448 | 0.43921 | 0.368599 |
| 19 | 45315445 | rs28399653 | G | A | **LUTHERAN** | **BCAM** | Combination Mutations | 0.01209677419 | 0 | 0.0131579 | 0 | 0 |
| 19 | 45315539 | rs3745159 | G | A | **LUTHERAN** | **BCAM** | Combinat<br>ion<br>Mutation<br>s | 0.02620<br>967742 | 0.0278 | 0.02875 | 0.0234<br>69 | 0.0215<br>2 |
| 19 | 45316704 | rs28399656 | T | A | **LUTHERAN** | **BCAM** | SNVs | 0.02769<br>3857 | 0.035 | 0.020025 | 0.0095<br>57 | 0.0094<br>85 |
| 19 | 45316804 | rs3810141 | C | T | **LUTHERAN** | **BCAM** | Combinat<br>ion<br>Mutation<br>s | 0.07703<br>927492 | 0.0682 | 0.1025 | 0.0879<br>96 | 0.0862<br>07 |
| 19 | 45316807 | rs3810140 | C | T | **LUTHERAN** | **BCAM** | Combinat<br>ion<br>Mutation<br>s | 0.07552<br>870091 | 0.0661 | 0.1025 | 0.0856<br>24 | 0.0841<br>9 |
| 19 | 45322316 | rs117737673 | C | T | **LUTHERAN** | **BCAM** | Combinat<br>ion<br>Mutation<br>s | 0.01560<br>926485 | 0.0095 | 0.02375 | 0.0222<br>28 | 0.0210<br>63 |
| 19 | 45322744 | rs1135062 | A | G | **LUTHERAN** | **BCAM** | Combinat<br>ion<br>Mutation<br>s | 0.330312<br>1853 | 0.2964 | 0.275 | 0.2753<br>29 | 0.2725<br>9 |
| 19 | 45322891 | rs28399658 | C | T | **LUTHERAN** | **BCAM** | Combinat<br>ion<br>Mutation<br>s | 0.03776<br>435045 | 0.0643 | 0.0575 | 0.0315<br>62 | 0.0296<br>53 |
| 19 | 45322962 | rs28399659 | A | T | **LUTHERAN** | **BCAM** | Combinat<br>ion<br>Mutation<br>s | 0.03850<br>050659 | 0.066 | 0.055625 | 0.0348<br>72 | 0.0297<br>57 |
| 19 | 49254504 | rs2071699 | G | A | **H** | **FUT1** | SNVs | 0.041792<br>54783 | 0.0462 | 0.065625 | 0.0578<br>46 | 0.0590<br>25 |
| 2 | 220080845 | rs60322991 | C | T | **LANGEREIS** | **ABCB6** | SNVs | 0.008611<br>95542 | 0.0201 | 0.00125 | 0.01911 | 0.0109<br>4 |
| 2 | 220081416 | rs57467915 | G | A | **LANGEREIS** | **ABCB6** | SNVs | 0.00405<br>2684904 | 0.0006 | 0.000625 | 0.0189<br>5 | 0.0087<br>83 |
| 22 | 43088971 | rs9623659 | C | T | **P1PK** | **A4GALT** | Combinat<br>ion<br>Mutation<br>s | 0.41683<br>46774 | 0.4468 | 0.340625 | 0.3523<br>91 | 0.3503<br>11 |
| 22 | 43089849 | rs11541159 | T | C | **P1PK** | **A4GALT** | SNVs | 0.41809<br>909 | 0.4485 | 0.341875 | 0.3867<br>2 | 0.3491<br>5 |
| 3 | 160804167 | rs2231257 | C | T | **GLOB** | **B3GALNT1** | SNVs | 0.04833<br>836858 | 0.0711 | 0.06 | 0.0456<br>55 | 0.0448<br>4 |
| 4 | 144920596 | rs7683365 | G | A | **MNS** | **GYPB** | Combinat<br>ion<br>Mutation<br>s | 0.33383<br>6858 | 0 | 0.360625 | 0.3456<br>92 | 0 |
| 4 | 89013496 | rs34783571 | C | T | **JUNIOR** | **ABCG2** | SNVs | 0.00860<br>3238866 | 0.0065 | 0.0175 | 0.0055<br>56 | 0.0052<br>04 |
| 4 | 89052323 | rs2231142 | G | T | **JUNIOR** | **ABCG2** | Combinat<br>ion<br>Mutation<br>s | 0.05186<br>304129 | 0.0517 | 0.07625 | 0.11801<br>7 | 0.1246<br>55 |
| 4 | 89061114 | rs2231137 | C | T | **JUNIOR** | **ABCG2** | Combinat<br>ion<br>Mutation<br>s | 0.106243<br>7059 | 0 | 0.11125 | 0.1075<br>81 | 0.11418<br>9 |
| 6 | 10586727 | rs56106312 | G | A | **I** | **GCNT2** | SNVs | 0.002517<br>623364 | 0.0043 | 0.0025 | 0 | 0 |
| 6 | 49580247 | rs16879498 | C | T | **RHAG** | **RHAG** | Combinat<br>ion<br>Mutation<br>s | 0.05945<br>121951 | 0 | 0.036875 | 0.0434<br>54 | 0.0420<br>61 |
| 7 | 142640004 | rs8176039 | T | C | **KELL** | **KEL** | Combinat<br>ion<br>Mutation<br>s | 0.02416<br>918429 | 0.0328 | 0.0075 | 0.0209<br>58 | 0.0173<br>51 |
| 7 | 142640113 | rs8176038 | A | G | **KELL** | **KEL** | SNVs | 0.011077<br>5428 | 0 | 0.001875 | 0.0083<br>55 | 0.0067<br>38 |
| 7 | 142640596 | rs8176036 | T | G | **KELL** | **KEL** | Combinat<br>ion<br>Mutation<br>s | 0.03373<br>615307 | 0.0274 | 0.02125 | 0.0528<br>12 | 0.0504<br>72 |
| 7 | 142640916 | rs8176034 | G | T | **KELL** | **KEL** | SNVs | 0.02668<br>680765 | 0 | 0.003125 | 0 | 0 |
| 7 | 142641752 | rs190890637 | G | A | **KELL** | **KEL** | SNVs | 0.001510<br>574018 | 0 | 0.0025 | 0.0002<br>16 | 0.0002<br>11 |
| 7 | 142651354 | rs8176059 | G | A | **KELL** | **KEL** | Combinat<br>ion<br>Mutation<br>s | 0.00352<br>4672709 | 0.0023 | 0.0025 | 0.0071<br>8 | 0.0069<br>76 |
| 7 | 142655008 | rs8176058 | G | A | **KELL** | **KEL** | Combinat<br>ion<br>Mutation<br>s | 0.04481<br>369587 | 0.0478 | 0.03125 | 0.0274<br>38 | 0.0268<br>98 |
| 7 | 30951658 | rs28362692 | C | T |  | **AQP1** | SNVs | 0.00805<br>6394763 | 0.004 | 0.005625 | 0.0264<br>68 | 0 |
| 7 | 30951664 | rs377506522 | A | G |  | **AQP1** | SNVs | 0.00201<br>4098691 | 0.0011 | 0.004375 | 0.0001<br>91 | 0.0001<br>99 |
| 9 | 136029645 | rs35898523 | G | T | **FORS** | **GBGT1** | SNVs | 0.09182<br>643794 | 0.1328 | 0.1 | 0 | 0 |
| 9 | 136037742 | rs2073924 | G | A | **FORS** | **GBGT1** | SNVs | 0.222557<br>9053 | 0.2193 | 0.232704 | 0.27771<br>1 | 0.11379<br>3 |
| 9 | 136136773 | rs8176696 | C | T | **ABO** | **ABO** | SNVs | 0.01359<br>516616 | 0 | 0.009375 | 0 | 0 |
| 1 | 43296572 | rs33954154 | C | T | **SCIANNA** | **ERMAP** | Combinat<br>ion<br>Mutation<br>s | 0.00856<br>8548387 | 0.0037 | 0.000625 | 0.0081<br>39 | 0.0063<br>98 |

**Table 2.**
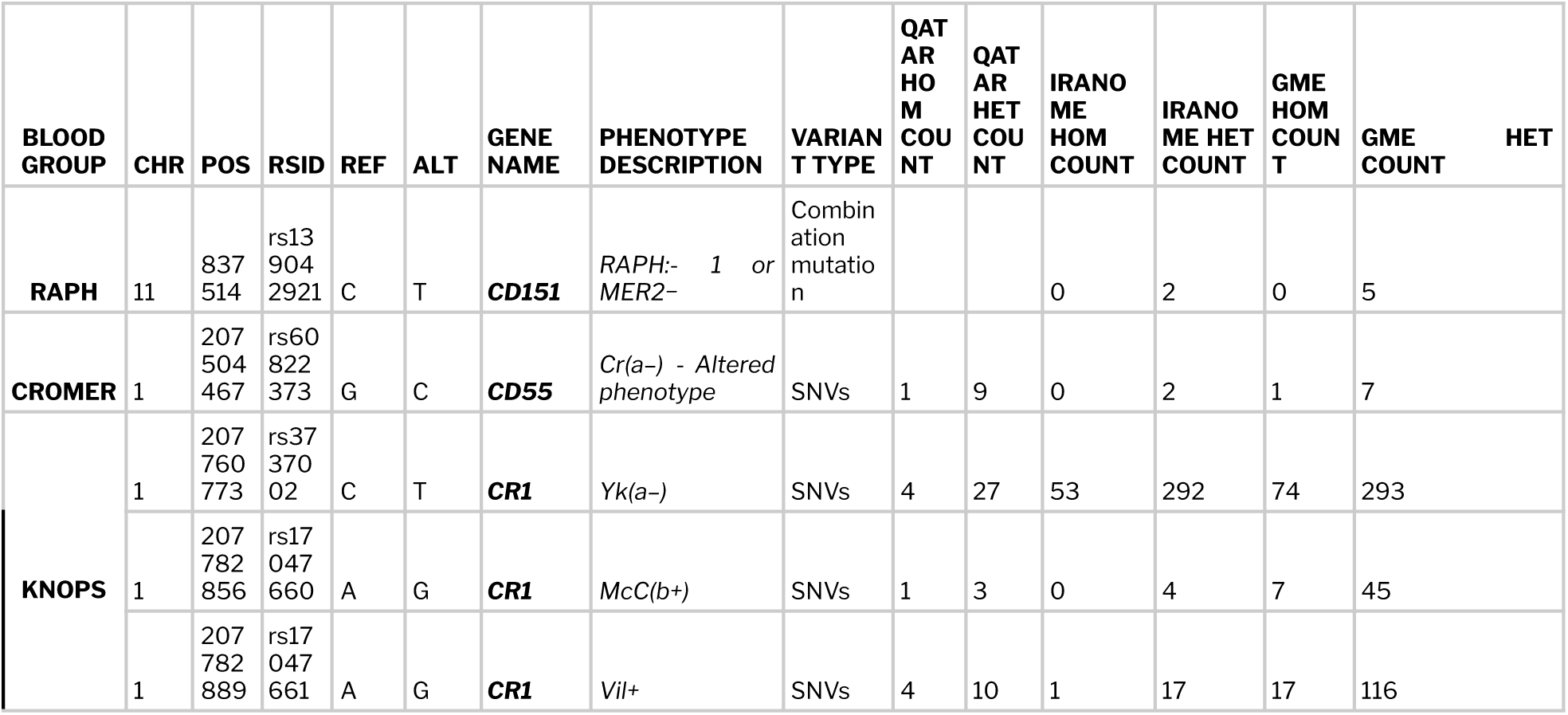
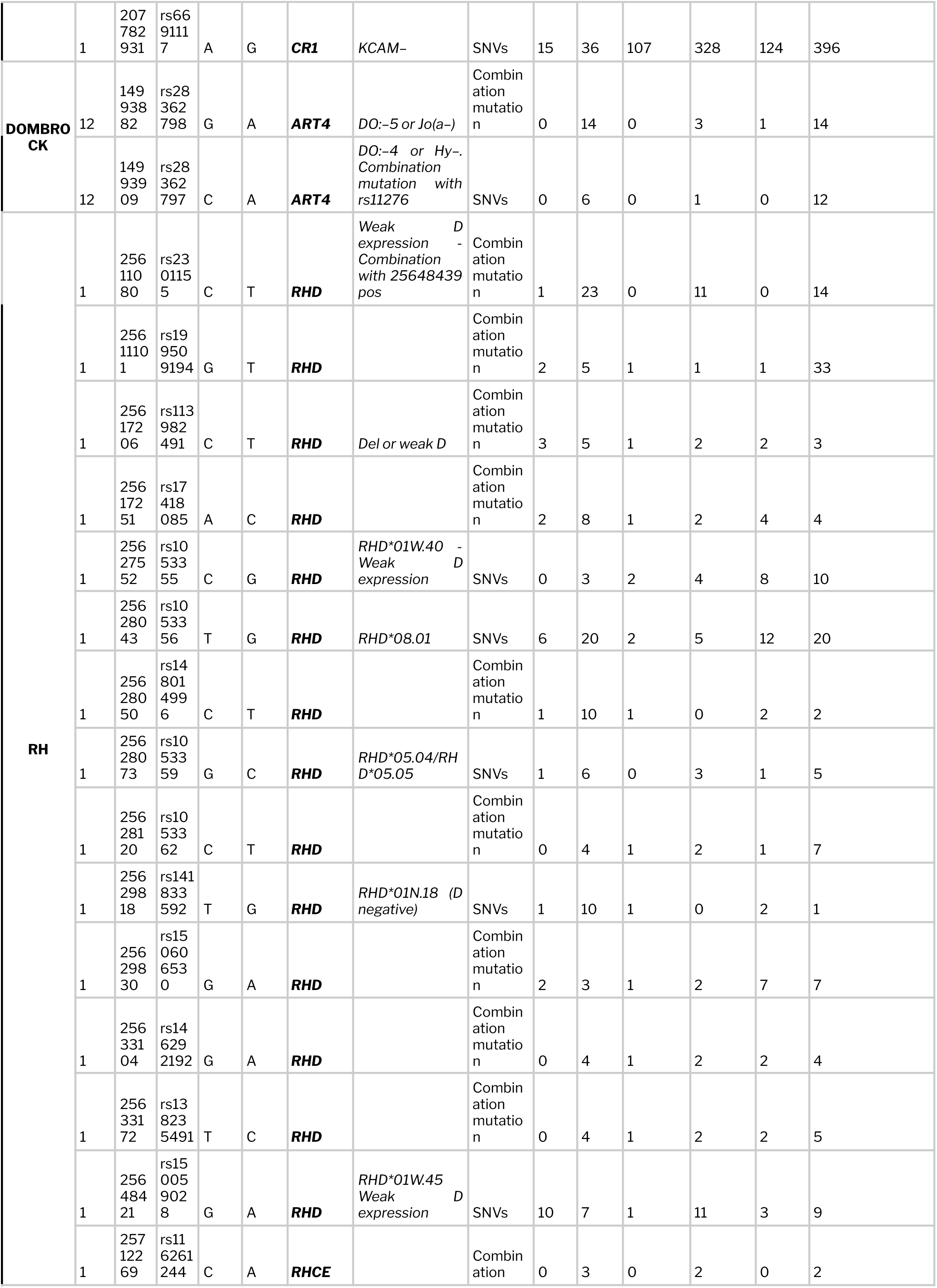
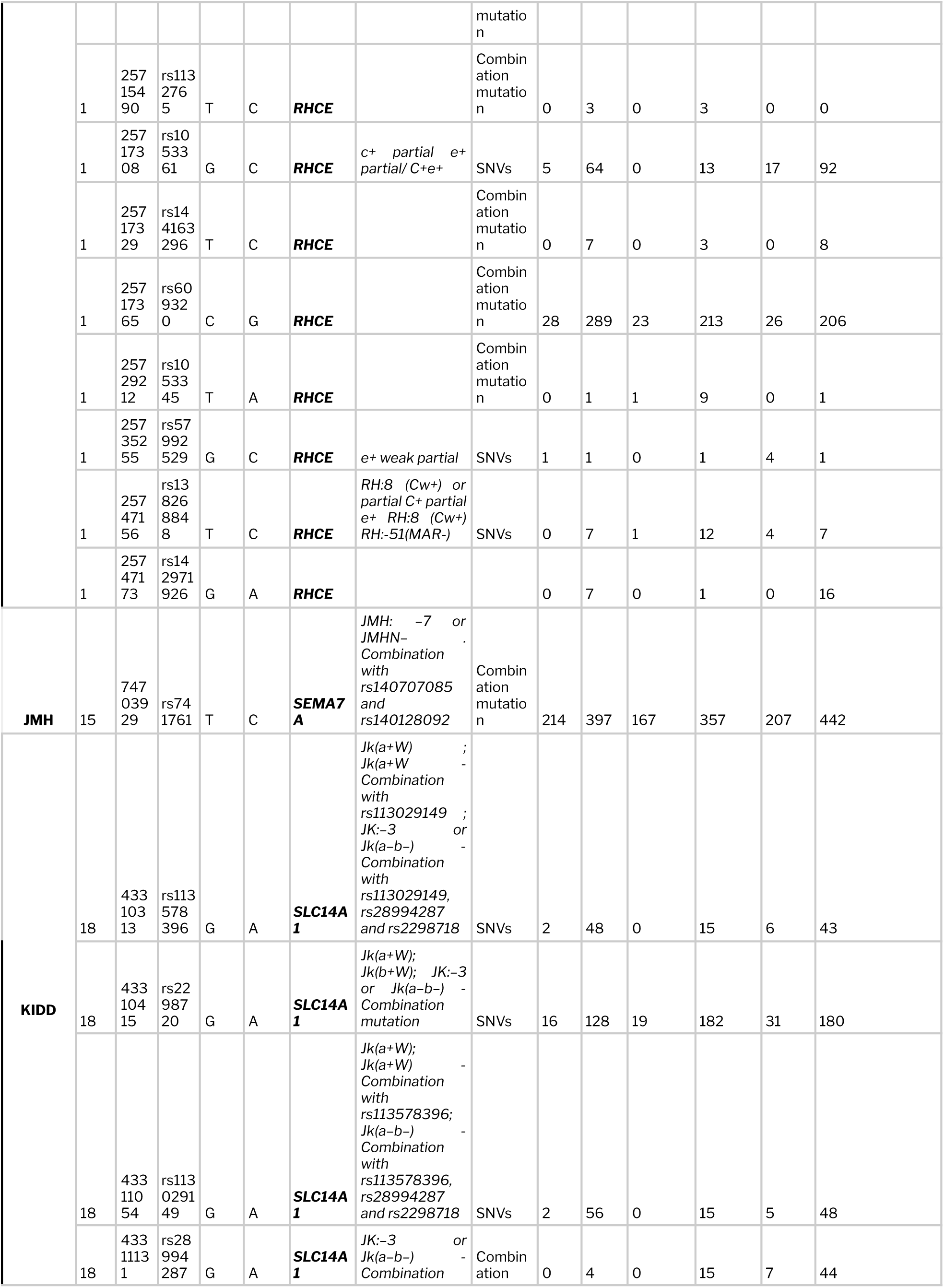
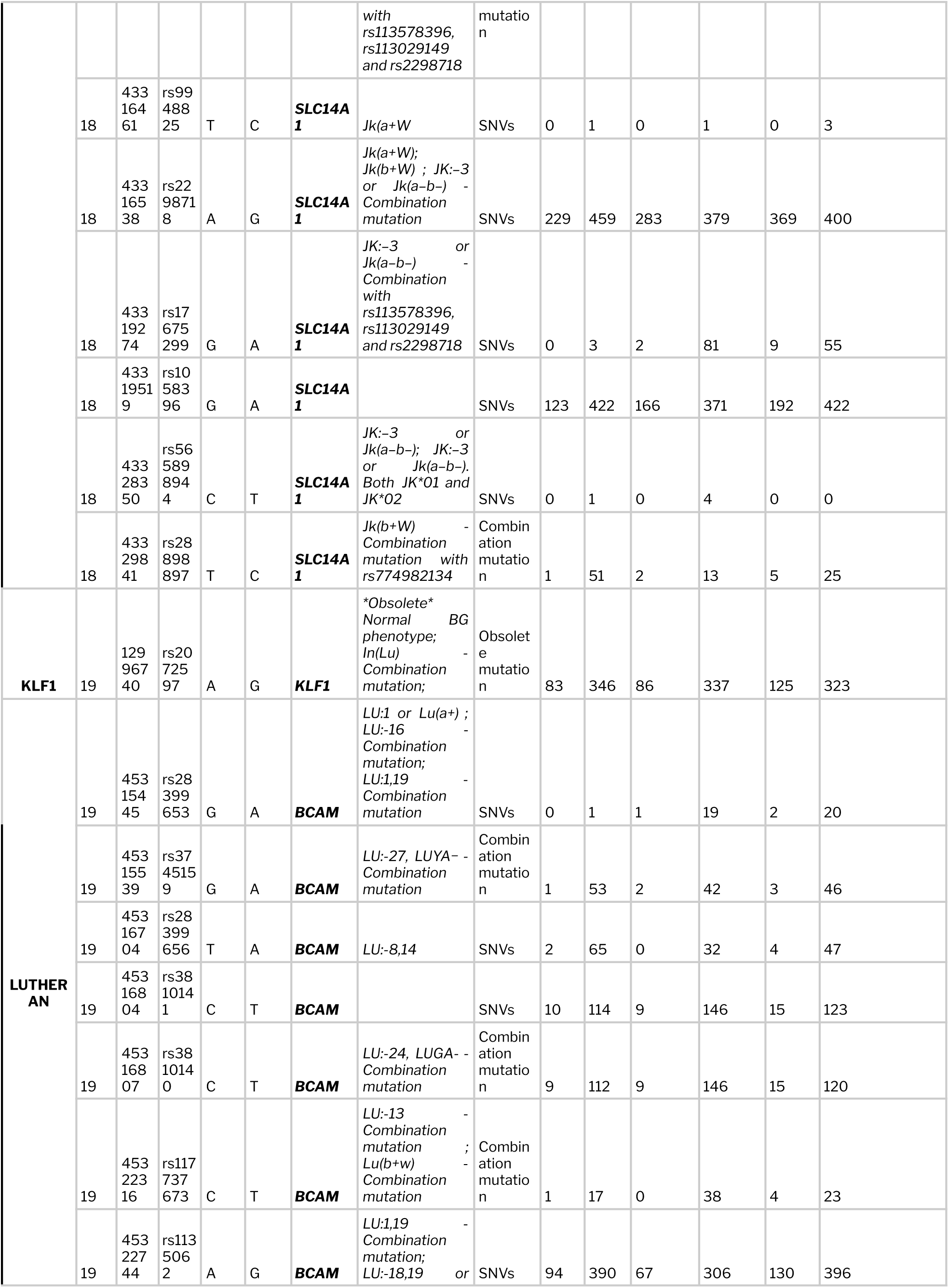
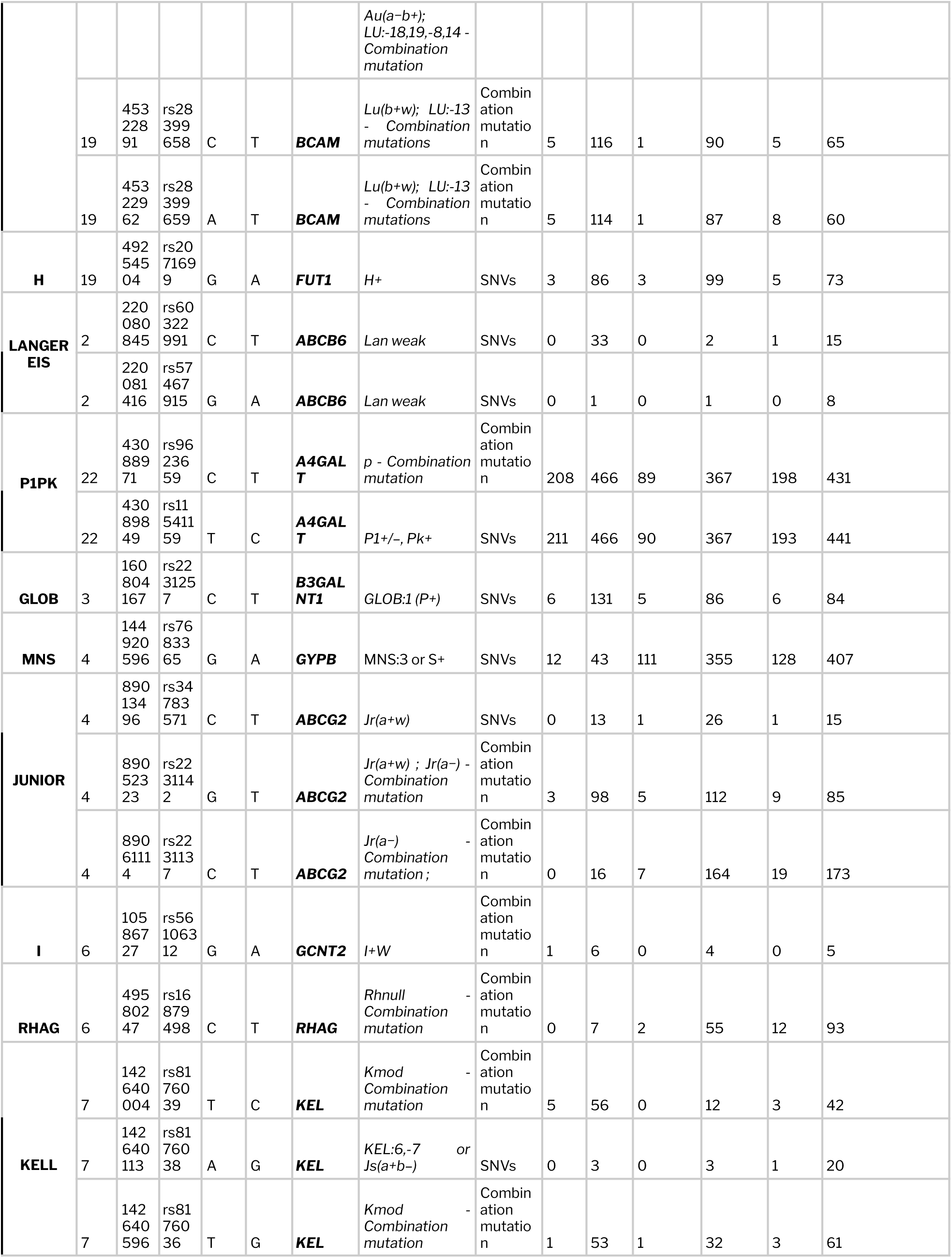
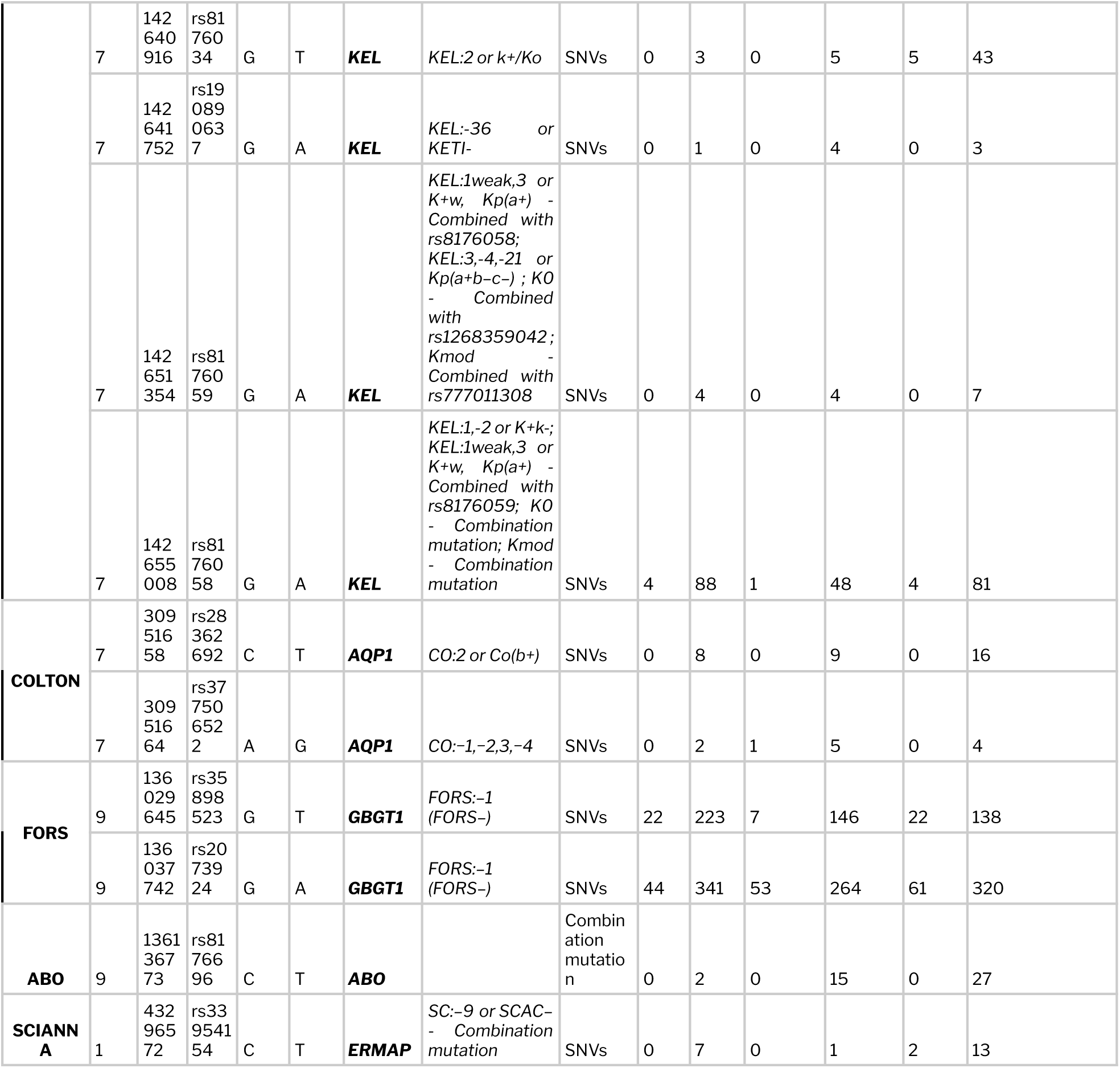
Summary of phenotypes of common blood group variants along with their corresponding zygosity information.

### Overview of potentially novel and rare blood group variants

Variants were systematically classified based on the dbSNP identifiers and were deemed as potentially novel if they lacked one. This analysis revealed a total of 1727 potentially novel unique variants across the datasets, of which 730 were found to span the exonic and splicing regions. Seventy (70) exonic variants which were not reported previously in global population frequencies including 1000 Genomes, gnomAD and EXAC were predicted to be deleterious by at least three or more computational tools. Distribution of these novel variants across different blood group genes along with their corresponding frequencies in the Middle Eastern datasets are summarized in **Supplementary Table 3.** A missense H blood group variant (p. Arg220Leu, chr19:49253880C>A) which was computationally predicted as deleterious, has a couple of alternate variants reported at the same position associated with weak H+ phenotype (p.Arg220Cys - FUT*01W.09 and p.Arg220His - FUT*01W.10). Yet another missense variant predicted from the same blood group (p. Try154Asp, chr19:49254079A>C) possess reported variants at the same amino acid position associated with weak and null H phenotypes (p.Tyr154Cys - FUT*01N.02, p.Tyr154His - FUT*01W.o5.01 and p.Tyr154Ter - FUT*01N.03).

Two missense variants belonging to JUNIOR blood group predicted as deleterious (p.Gly262Glu, chr4:89039317C>T and p.Arg246Leu, chr4:89039365C>A) were found to occur at the same amino acid sites which were reported to be associated with Jr(a-) phenotypes (p.Gly262X - ABCG2*01N.05 and p.Arg246X - ABCG2*01N.03 respectively).

### Distribution of null and partial/weak antigens

Weak or partial expression of RBC antigens carries increased risk of hemolytic transfusion reactions as they are often mistyped as antigen-negative. Of the total variants analysed, our study reports 19 variants responsible for weak/partial or null blood group phenotypes. The RHD*weak D type 4.0 allele *(rs1053355)* was observed in 10 individuals in a homozygous state. This allele, responsible for the weak expression of D antigen, was found to be often mistyped as D- in serological tests (42). In addition, one of the rare low frequency antigens of the RH blood group system, C^w+^ (Rh8 - rs138268848) was found in about 0.17% of Middle Eastern populations (0.13% in Iranome and 0.36% in GME). This antigen has been reported to have serious transfusion implications including hemolytic disease of the newborn when mismatched (43).

A weak JK*A allele (JK*01W.01 - rs2298720) of the KIDD blood group system, associated with weakened expression of Jk^a^ is found capable of causing alloimmunization when mistyped serologically as Jk_null_ (44), (45). Distribution of this variant is found to vary highly among various global populations with an allele frequency as high as 40.6% in East Asians, 26.8% in South Asians, 18-19% in Latino/Admixed Americans and African/African Americans, 12% in Europeans of Finnish origin, 10% in Ashkenazi Jews to as low as 7% in Middle Easterners, 6% in Europeans of non-Finnish origins and 3% in Amish population. **Supplementary Table 4** enlists the complete details of weak/partial alleles identified in the study dataset.

### Clinically significant blood group variants

The KELL blood group system is significant in the field of transfusion medicine owing to the severe transfusion incompatible reactions induced by its antigens (46). This system comprises over 20 different antigens, of which K1 (K) has been well studied as a strong immunogen capable of causing hemolytic disease of the newborn in sensitized mothers. Approximately 9% of the random RBC donor samples were found to react against K1 (46,47). Anti-K1 is ranked third most clinically significant antibody after ABO and RH(D). HDN induced by anti-K is characterized by immune destruction of K+ erythroid (47) progenitor cells by macrophages in the fetal liver (48). Frequency of this variant in Middle Easterners was found to be 3.5% which was comparable to Europeans of non-Finnish origin (4.1%) and differ from South Asians (<1%) (49), (50)

Anti-Cr^a^ is a rarely encountered antibody against the Cr(a) antigen of the CROMER blood group system. CROM:c.679G>C (rs60822372) variation in exon 6 of CROM gene is responsible for Cr(a-) phenotype which is characterized by the absence of Cr(a) antigen on the RBC membrane. Studies have shown negative crossmatching of individuals carrying the variant in homozygous state and positive while in heterozygous state (51). This variant was observed at a frequency of about 10% in Middle Eastern populations which was found to be comparable to African/African Americans and South Asians (52), (53),(54).

### Blood group phenotyping of Qatari subpopulations

The genomic data of 1005 Qataris comprises 5 major subpopulations namely African (n=70), Arabian (n=193), Bedouin (n=490), Persian (n=170) and South Asian (n=76). Distribution of blood group related variants in different subpopulations was identified and corresponding phenotypes were predicted . In addition, systematic comparisons of allele and phenotype frequencies among the subpopulations were performed.

### Comparison and statistical analysis of blood group variants/profiles among Middle Eastern population datasets

Distinct differences in the minor allele frequencies of blood group alleles between the Middle Easterners and various other global populations were statistically estimated using Fisher’s exact test. A total of 26 variants mapping back to 12 blood group systems were found statistically distinct in all the Middle Easterners (Qatar, GME and Iranome) in comparison to global datasets (1000 Genomes Project and gnomAD version?). Summary of these variants along with their p-value is provided in **Supplementary Table 5a.** In addition, differences in the pattern of distribution of these blood group variants were checked among the 5 major Qatari subpopulations. The Arabian subpopulation was found to carry the maximum number of variants whose frequencies were statistically distinct from the overall frequency in the Qatar dataset. Complete details of these variants are tabulated in **Supplementary Table 5b.**

### User friendly data access

alnasab (*Alleles and antigens in Arab and Persian populations associated with blood groups)* is a user-friendly online search engine, composed of a comprehensive collection of blood group related variations mined from various population scale datasets of the Greater Middle Eastern region. The resource encompasses a total of 2149 blood group related variants fetched from about 47 blood group genes. The search interface enables the user to query the database based on gene name, variant name (chr-pos:ref>alt), nucleotide change and protein change. Variant information is provided in six different sections namely, (i) Blood group system, (ii) Variant details, (iii) Transcript details, (iv) Variant annotation, (v) Global allele frequencies and (vi) Allele frequencies - Greater Middle East. **Figure 4a and 4b** illustrate the search features and various sections of result display of the resource.

**Figure 4.**
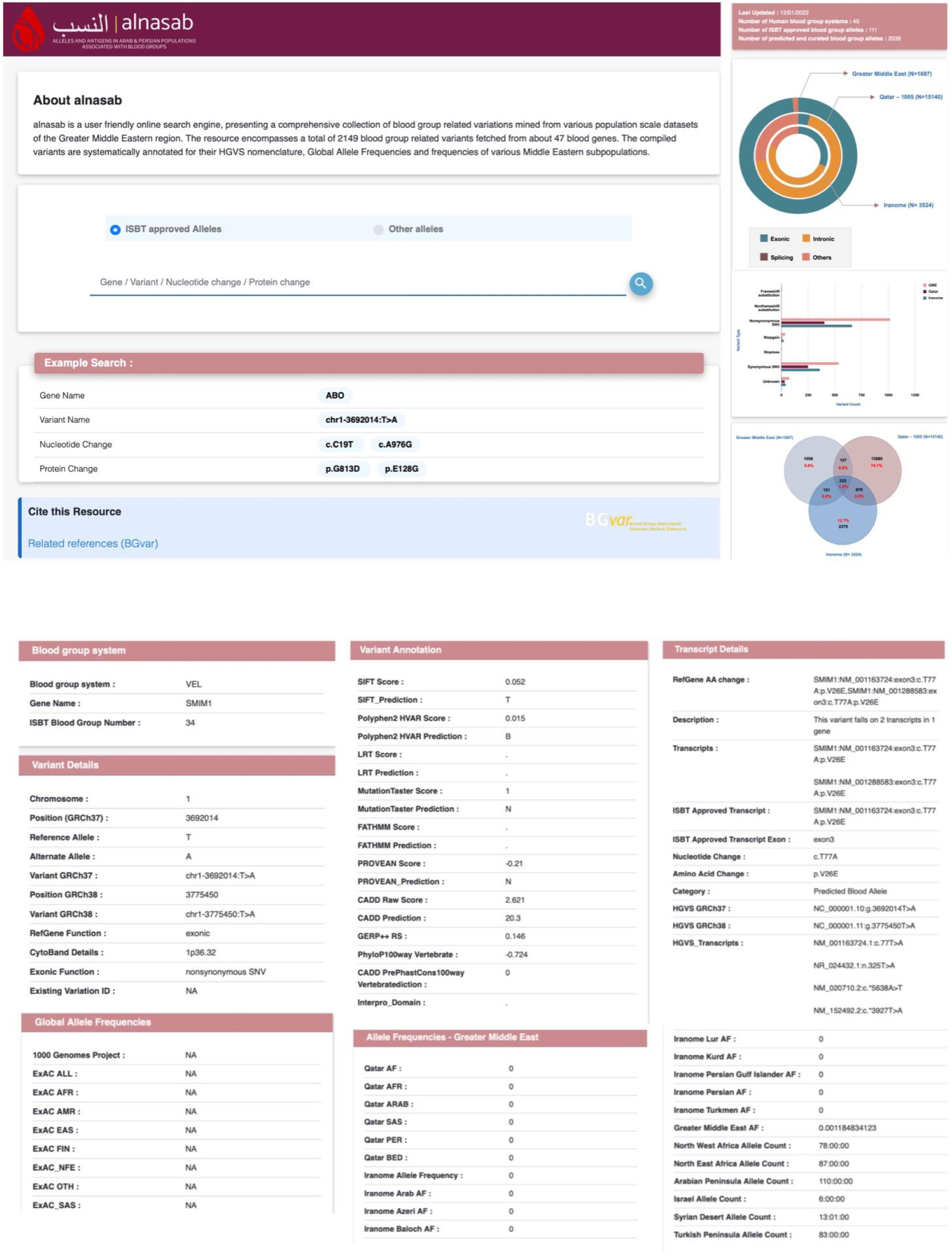
Search query and sections of result display in alnasab. (A) Homepage of the web resource with query search examples. (B) Result display which provides an extensive summary of variant functional annotations in various sections

## Discussions

Genomic analysis presented in this study provides the blood group genotype profiles of the Middle Eastern populations utilizing the genome sequencing datasets from Greater Middle East Variome, The Qatar genome and the Iranome. The study reports an array of weak, partial, null and putative novel and rare antigens which were predicted to have potential impact on the functionality. The analyzed and filtered blood group variants along with extensive functional annotations and allele frequencies are provided as a comprehensive online repository - **alnasab,** *Alleles and antigens in Arab and Persian populations associated with blood groups.* The resource is accessible at https://clingen.igib.res.in/alnasab/

### Clinical impact

An extended screening of blood group antigens between the donors and the recipients other than the major ABO, RH groups and minor DUFFY, KELL and KIDD groups is often triggered only after encountering clinical complications. Frequencies of a total of 26 variants belonging to 12 different blood group systems were found to differ significantly from other global populations. Fetal anaemia and Hemolytic disease of the fetus and newborn (HDFN) are the most serious consequences of transfusion in the majority of pregnancy settings (55). The clinically significant antibodies involved in such cases include Rh, C, E, c and e antigens with greater than 50% risk of mild to severe HDFN (56),(57). Our study reports a weak RHD allele (*RHD*weak D type 4.0*) with an overall frequency of 0.01% in the Middle eastern population. Often patients carrying this allele get serologically categorized to be transfused with D-RBCs. A recent report from a transfusion work group recommendation explained the necessity of RHD molecular genotyping in expectant mothers which would help categorize RHD alleles that can further allow safe D+ transfusion (42).

### Literature comparison

There exists a handful of research studies and reports documenting the blood group profiles of donors across various parts of the Arabian peninsula. Phenotypes of ABO, RH and various other minor blood group systems predicted by standard serological methods were found to match with the genotyping profiling results of the study. (50), (58), (58,59), (60), (61). Js^a^ (KEL6) is a low-incidence antigen reported in less than 1% of the general global populations, whereas on other hand, could be prevalent upto 19.5% in African Americans (62). Js(a+b-) phenotype which was not previously reported in serological studies was found in one sample from North East Africa. In compliance with earlier evidence, frequencies of minor blood group phenotypes including Fy(a+b+), K-k+, Lu(a-b+), Jk(a+b+), Do(a+b+), Co(a+) were found comparable.

### Limitations and future needs

Owing to the limitations in the available public datasets, our study is mainly limited by the fact that RBC antigenic expressions regulated by large deletions and insertions (especially in RH and MNS blood groups) have not been profiled. In addition, the level of concordance with serology based phenotype predictions to investigate the functional implications of novel and rare variants remains to be explored and validated in future studies.

## Conclusion

Middle Easterners represent one of the most genetically diverse populations with increased prevalence of genetic disorders owing to their consanguinity. Autosomal recessive genetic blood disorders including sickle cell disease (SCD), beta thalassemia and hemoglobinopathies are found to be the most common in the Middle Eastern population (63), (18). Increased burden of anemia in women and children is also reported in this population ranging between 22.6-63% in pregnant women, 27-69.6% in women of reproductive age and 23.8-83.5% in children under age of 5 (64). The overall prevalence rates per 1000 population for beta thalassemia and SCD were found as 13.6 and 49.6 respectively (19). Severe forms of such disorders and other chronic illnesses require frequent blood transfusions often leading to dysfunction of multiple organs, if mismatched

(65). Extensive population scale characterization of blood group antigen profiles can significantly increase the clinical outcomes of transfusion practices. Numerous national initiatives including Makkah Region Quality Program (MRQP), Central Board of Accreditation for Health Institutions (CBAHI), The Saudi Food and Drug Authority (SFDA), Western Region Transfusion Medicine Group and Saudi Society of Transfusion Medicine (SSTM) have been organized by the Ministry of Health of Saudi Arabia which aims at improving the blood transfusion services in the region (66). Our study showcases the utility of population scale genome sequencing datasets in elucidating the complete blood antigen profiles of the population to guide and improve transfusion practices and outcomes.

## Data Availability

All data produced in the present work are contained in the manuscript

https://clingen.igib.res.in/alnasab/

## Acknowledgements

VS conceived and designed the project. MR and VS contributed in writing the manuscript. KP designed the database. All authors approved the final manuscript. Authors acknowledge funding from CSIR India. The funders had no role in the preparation of the manuscript or decision to publish. The authors acknowledge the constructive suggestions given by Srashti Jyoti Agrawal and Vishu Gupta.

## Supplementary Datasets

**Supplementary Table 1.** Brief tabulation of number of variants found associated with human blood group genes in the datasets used in the study.

[Summary of blood group variants in study datasets](https://docs.google.com/spreadsheets/d/18738idgnMrDd34gXghSkBE0x9eK14GmAg_J8B4bu3cE/edit?usp=sharing)

**Supplementary Table 2.** Tabulation of blood group alleles predicted to match ISBT approved phenotypes and reference genome nomenclature in Middle Eastern population datasets

[Predicted phenotypes of blood group systems in Middle Eastern population](https://docs.google.com/spreadsheets/d/18738idgnMrDd34gXghSkBE0x9eK14GmAg_J8B4bu3cE/edit?usp=sharing)

**Supplementary Table 3.** Summary of distribution of novel and rare blood group variants in GME datasets predicted to be deleterious by at least three or more computational tools.

[Tabulation of potentially novel and deleterious blood group variants in GME datasets](https://docs.google.com/spreadsheets/d/18738idgnMrDd34gXghSkBE0x9eK14GmAg_J8B4bu3cE/edit?usp=sharing)

**Supplementary Table 4.** Comprehensive tabulation of weak/partial or null allele variants found in Middle Eastern populations along with their zygosity information.

[Summary of weak or partial antigens identified in the study dataset](https://docs.google.com/spreadsheets/d/18738idgnMrDd34gXghSkBE0x9eK14GmAg_J8B4bu3cE/edit#gid=0)

**Supplementary Table 5a.** Summary of statistically distinct blood group variants identified between the Middle East and global population datasets

[Distinct blood group variants between the Middle East and global populations](https://docs.google.com/spreadsheets/d/18738idgnMrDd34gXghSkBE0x9eK14GmAg_J8B4bu3cE/edit#gid=0)

**Supplementary Table 5b.** Summary of statistically distinct blood group variants observed among the Qatari subpopulations

[Distinct blood group variants among Qatari subpopulations](https://docs.google.com/spreadsheets/d/18738idgnMrDd34gXghSkBE0x9eK14GmAg_J8B4bu3cE/edit#gid=0)

**Supplementary Figure 1**. Distribution of statistically distinct blood group variants in various Middle Eastern datasets and among Qatari subpopulations

**Supplementary Figure 1.**
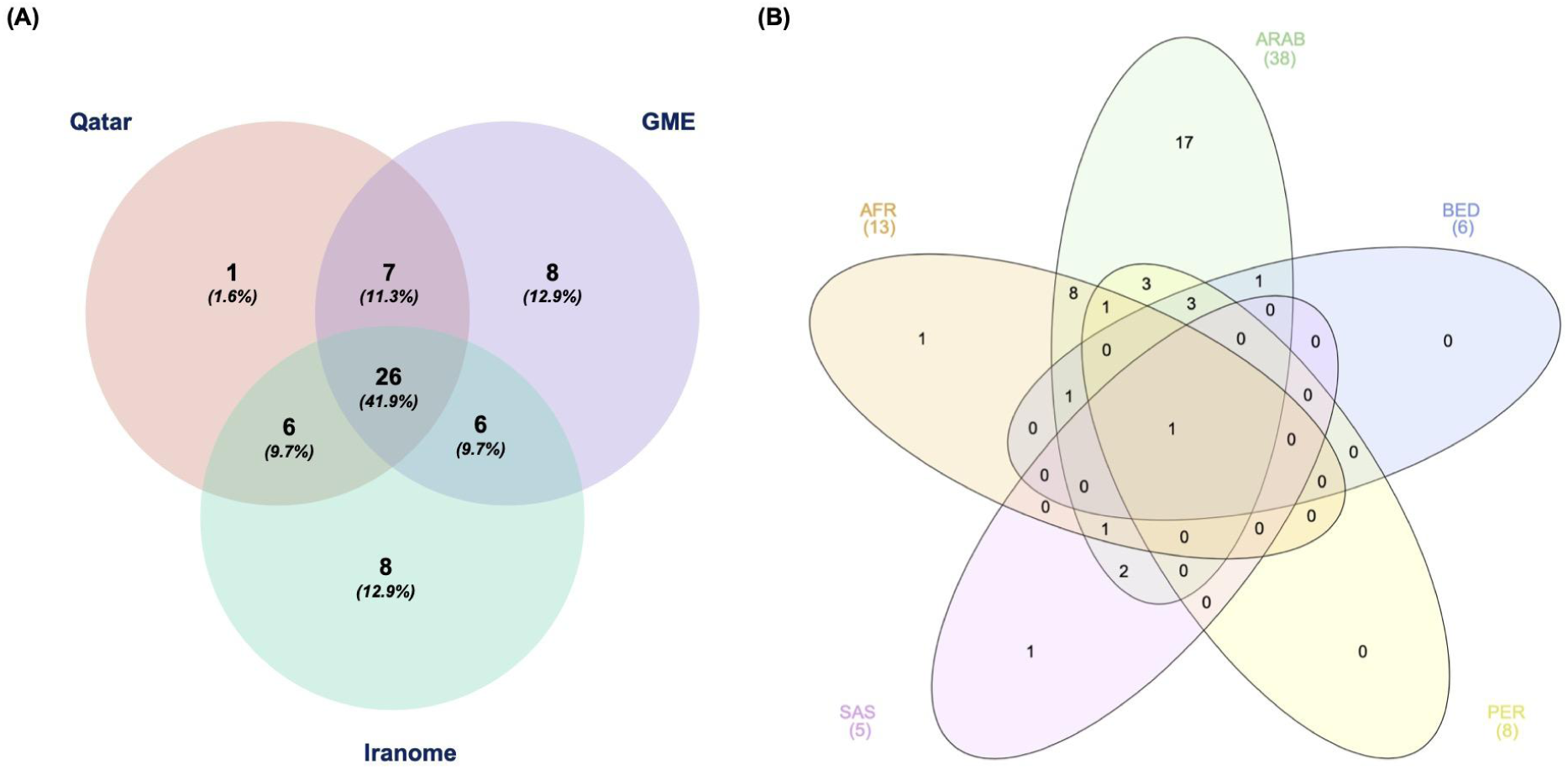
Distribution of statistically distinct blood group variants. (A) Summary of blood group variants which were found statistically distinct in various Middle Eastern datasets in comparison to global population data. (B) Distribution of statistically distinct blood group variants among the major Qatari subpopulations in comparison to the overall Qatar frequencies.

